# FEIBA® Reversal for Apixaban and Rivaroxaban in Patients with Intracranial and Non-intracranial Hemorrhages

**DOI:** 10.1101/2020.08.19.20178061

**Authors:** Aleah R. Hunt, Shawn N. Coffeen, Dane L. Shiltz, Calvin Ice, Jessica Parker

## Abstract

**Background:** Since FDA approvals, apixaban and rivaroxaban use has steadily increased. Currently, FEIBA® has been used off-label for factor Xa inhibitor reversal yet there are limited studies to support this practice. Therefore, additional safety and effectiveness data is needed for apixaban and rivaroxaban reversal in patients with an associated bleeding event.

**Methods:** The following retrospective study evaluated patients who received at least one dose of FIEBA® for the reversal of apixaban or rivaroxaban. One hundred forty-seven patients with an acute bleed were evaluated. The primary study outcome sought to determine the percentage of patients who achieved excellent or good hemostatic effectiveness within 12 hours of FEIBA® administration. The primary safety outcomes assessed the percent of patients who experienced an inpatient adverse event defined by thrombosis or mortality during hospital admission post-FEIBA® administration.

**Results:** Among the 147 patients evaluated, 58 experienced an intracranial hemorrhage (ICH) and 89 experienced a non-ICH bleeding event. One hundred fifteen patients (78%) achieved excellent or good hemostasis. Three patients (2%) experienced a thrombotic complication while another 3 patients (2%) had a hemorrhagic complication. A total of 15 patients (10%) experienced in-hospital mortality following FEIBA^®^ administration.

**Conclusion:** This retrospective study supports the safety and effectiveness of FEIBA® for the management of acute bleeding events secondary to apixaban and rivaroxaban. FEIBA® achieved excellent or good (collectively defined as effective) hemostasis similarly for ICH and non-ICH bleeding events in patients receiving apixaban or rivaroxaban. Furthermore, the thromboembolism outcomes associated with FEIBA® were minimal. The effectiveness and safety outcomes support FEIBA® as a plausible apixaban and rivaroxaban reversal agent, notably among patients experiencing ICH.

## Background

Apixaban and rivaroxaban use has steadily increased relative to warfarin for the management of multiple cardiovascular indications.^1^ Anticoagulants are commonly used for the treatment of venous thromboembolism or for the prevention of cardioembolic stroke or systemic embolism secondary to atrial fibrillation.^1^ The non-vitamin K antagonist oral anticoagulants (NOACs), apixaban and rivaroxaban, selectively and reversibly inhibit clotting factor Xa, a major component in the blood coagulation cascade common pathway. This mechanism of action results in the inhibition of platelet aggregation through decreased thrombin (factor IIa) generation. Several advantages exist for the use of NOACs over the traditional vitamin K antagonist oral anticoagulant, warfarin. This includes minimal monitoring parameters as well as fewer drug-drug and drug-food interactions.^1,2^ Consistent, therapeutic anticoagulation is achieved once a NOAC reaches steady state without the routine need to monitor blood concentrations. Compared to warfarin, the relative consistency of apixaban and rivaroxaban blood concentrations reduce the risk for thrombotic and hemorrhagic complications.

Patients receiving anticoagulants, including NOACs, have a significant risk for bleeding following trauma, emergency surgery, and fall among other insults. In an emergent situation, a proper reversal agent can reduce blood loss, restore hemodynamic stability, and reduce mortality. In 2018, the United States Food and Drug Administration (FDA) approved the first reversal agent, andexanet alfa, for apixaban and rivaroxaban.^2,3^ Prior to the approval of andexanet alfa, health care providers used human prothrombin complex concentration (Kcentra^®^), recombinant coagulation factor VIIa (NOVOSEVEN RT®), alternative 3-and 4-factor prothrombin complex concentrates (PCCs), or idarucizumab (Praxbind®) to reverse select NOACs and other anticoagulants.^1^ However, the study promoting andexanet alfa approval revealed noteworthy limitations including the small population size and no control group.^3,4^ Furthermore, andexanet alpha ranks atop the drug product cost hierarchy for anticoagulant reversal. Given the preexisting andexanet alpha literature, notable expense tied to its use, and availability of less costly alternative coagulation factors that may prove at least as effective, investigating the outcomes of other anticoagulant reversal agents remains prudent.

Factor eight inhibitor bypass activity (FEIBA^®^), an activated prothrombin complex concentrate (aPCC), was first FDA approved for the treatment of hemophilia in 1986.^6,7^ More recently FIEBA^®^ has been used as a reversal agent for warfarin^6^ yet remains less studied for the reversal of NOAC therapy, specifically apixaban and rivaroxaban. The following retrospective study evaluated FEIBA^®^ for the treatment of a significant bleeding event in patient receiving apixaban or rivaroxaban. Using the safety and efficacy outcomes outlined in the ANNEXA-4 study^3^, FEIBA® was evaluated for the reversal of apixaban-and rivaroxaban-attributed intracranial (ICH) and non-intracranial (non-ICH) bleeding events.

## Methods

### Trial Design

This Spectrum Health institutional review board (IRB)-approved retrospective study evaluated the clinical outcomes among patients who received at least one dose of FIEBA® to reverse apixaban or rivaroxaban. Patients were enrolled from January 1, 2014 to June 1, 2019. FEIBA® safety and effectiveness for apixaban and rivaroxaban reversal was established via electronic medical record review.

The primary study objective sought to evaluate the effectiveness and safety of FEIBA® for the reversal of apixaban and rivaroxaban. The primary hypothesis proposed that FEIBA^®^ is a safe, effective, and cost-beneficial alternative for patients needing apixaban or rivaroxaban reversal following an ICH or non-ICH bleeding event.

### Patient selection

Key inclusion criteria included the following: receipt of at least one dose of FEIBA® administered in the emergency department or inpatient setting at Spectrum Health - Butterworth Hospital, Blodgett Hospital, or specific affiliate hospital (in order to verify dose and administration time); age 18 years or older; and therapeutic anticoagulation with total daily dose ranges of apixaban 5-20 mg or rivaroxaban 15-30 mg prior to FEIBA® administration. Key exclusion criteria included vulnerable populations (pregnant, breastfeeding, or incarcerated subjects); FEIBA® administration for non-apixaban or rivaroxaban reversal; anticipated subtherapeutic anticoagulant effect (known administration of apixaban or rivaroxaban ≥48 hours prior to FEIBA® administration); inability to determine anticoagulant use prior to admission and history of thrombosis (myocardial infection, pulmonary embolism, deep vein thrombosis, and/or arterial thrombosis) within the past 3 months. The totals and reasons for exclusions are detailed in Appendix A.

Additionally, a subgroup analysis of patients who would have been candidates to receive andexanet alfa instead of FEIBA was performed. To identify this population, additional exclusion criteria from the ANNEXA-4 study^3^ were further applied to the study population. Patients were excluded in this subgroup analysis if they had a Glasgow Coma Score (GCS) < 7, admitted to surgical services (trauma, general surgery, cardiac surgery, neurosurgery/neurocritical care, other surgical service, cardiology, internal medicine, medical ICU, other admitting service), indication of apixaban or rivaroxaban reversal for emergency surgery unrelated to a bleeding event, received FFP, or received other coagulation factor products.

### Outcome Measures

The primary effectiveness outcome assessed the percentage of patients who achieved excellent or good hemostasis within 12 hours of FEIBA® administration (see supplemental Appendix B and Table 1 for hemostasis definitions). The primary safety outcome determined the percent of patients who experienced a post-FEIBA® inpatient adverse outcome which included thrombosis or mortality during hospital admission. Serum hemoglobin, computed tomography (CT) results, and number of packed red blood cells or fresh frozen plasma products were evaluated to determine the effectiveness of FEIBA^®^ reversal.

### Statistical Analysis

Normally distributed numeric data were expressed as mean ± standard deviation and analyzed via two sample independent t-test. Non-normally distributed numeric data were expressed as median [25 ^th^, 75 ^th^ percentile] and analyzed via Wilcoxon Rank Sum. Categorical data was expressed as frequency (percent) and analyzed via Chi-Square or Fishers Exact Test (denoted by asterisk on p-value).

## Results

One hundred forty-seven patients were evaluated for a bleeding event (Table 3). Eighty-nine patients (61%) experienced a non-ICH and 58 patients (39%) experienced an ICH.

### Primary outcome

Overall, 115 patients (78.2%) achieved excellent or good hemostasis (collectively defined as effective hemostasis). Among the 89 and 58 patients experiencing non-ICH and ICH events, respectively, 46 patients (51.7%) and 43 (74.1%) specifically achieved excellent hemostasis. Compared to non-ICH bleeding events, there was a statistically significant association between excellent hemostasis in patients experiencing an ICH (51.7% vs. 74.1%, respectively; p=0.0049). However, there was no statistically significant difference observed between ICH and non-ICH events when excellent and good hemostasis outcomes were combined (81% vs. 76.4%, respectively; p=0.5061). Effectiveness details are provided in Table 4.

### Safety outcomes

Among the 147 patients, 3 (2.0%) experienced a thrombotic complication - 2 patients (2.2%) in the non-ICH group and 1 (1.7%) in the ICH group. For hemorrhagic complications, 3 patients (2.0%) in the full cohort had a hemorrhagic event with 2 patients (2.2%) in the non-ICH and one patient (1.7%) in the ICH groups. Fifteen patients expired after FEIBA® administration - 9 patients (10.1%) patients in the non-ICH group and 6 patients (10.3%) in the ICH group. While there was no difference in the total median FEIBA® exposure (p=0.5776), there was a statistically significant difference in the weight-based median FEIBA^®^ exposure (p=0.0155). However, no statistically significant clinical safety outcomes were observed. Tables 2 and 4 reflect FEIBA^®^ dosing and clinical safety outcomes, respectively.

### Subgroup analysis

Applying the ANNEXA-4 study inclusion criteria^3^, 97 (65.9%) of the 147 patients would have been candidates to receive andexanet alfa. Among these 97 patients, 80 (82.5%) achieved excellent or good hemostasis while 1 patient (1%) experienced a thrombotic complication and 9 patients (9.3%) expired after FEIBA® administration. No statistically significant differences in effectiveness or safety were observed. The outcomes from this specific cohort are provided in Table 5.

## Discussion

### Limitations

Due to the retrospective design, abstracting complete data from the electronic medical record occasionally proved challenging given the reliance upon adequate documentation. Despite utilizing established hemostasis criteria for different bleeding events^3^, certain cases lacked the necessary documentation to ascertain achievement of hemodynamic stability or last time of NOAC administration. Consequently, these cases were excluded from the study. Furthermore, inconsistent documentation prior to admission prevented a consistent determination of last apixaban or rivaroxaban administration, complicating a determination of FEIBA intent for anticoagulant reversal or for other, albeit unlikely, indications. Patients were included if they received any dose of FEIBA.

Each patient’s dose of FEIBA® was evaluated to ensure clinically appropriate reversal. Also, the amount of blood products given varied between patients, potentially skewing our results regarding the percent change of hemoglobin from baseline after FEIBA^®^ administration. Lastly, this study was single arm similar to the ANNEXA-4 trial.^3^

### Conclusion

The results support the effectiveness of FEIBA® to achieve excellent or good hemostasis in patients receiving apixaban or rivaroxaban. FEIBA^®^ collectively achieved effective hemostasis for both ICH and non-ICH populations, similar to the outcomes observed in the ANNEXA-4 trial. The safety results further demonstrate that thrombotic and hemorrhagic complications were reduced compared to the rate observed in the ANNEXA-4 trial for andexanet alfa used for apixaban and rivaroxaban reversal.^3^ Overall, FEIBA® achieved effective hemostasis combined with favorable thrombotic and hemorrhagic risks showing that FEIBA^®^ could be viewed as a potential NOAC reversal agent, notably for patients presenting with ICH.

In addition to an effective reversal agent, FEIBA^®^ appears to offer a cost-effective alternative to andexanet alfa. The FEIBA® dose of 50 units/kg, a weight-based dose arbitrarily classified as high-dose and similar to the median 48.7 units/kg dose used in this retrospective study^8^, corresponds to a cost of $116 USD per kilogram of actual body weight or nearly $10,600 per dose based on the 91.3 kg median body weight observed in this study.

Furthermore, only 3 patients (2%) received a second FEIBA® dose. Comparatively, the cost associated with andexanet alfa is at least $24,750 based on the package insert dosing recommendation.^9^ The subgroup analysis further demonstrated that FEIBA^®^ was associated with increased safety and similar effectiveness to andexanet alfa.^3^ Additional prospective, randomized controlled trials comparing FEIBA^®^ with andexanet alfa should be performed to support the association observed in this retrospective study.

## Data Availability

The data included in the manuscript has been deidentified and retained on an internal redCAP database.

## Supplemental Material

**Table 1.** Reversal indication and ICH breakdown

| Intracranial hemorrhage | Indication for reversal | Frequency | Percent |
| --- | --- | --- | --- |
| No | Trauma (non-ICH) | 25 | 17.01 |
| No | Bleeding event (non-ICH) | 37 | 25.17 |
| No | Emergency surgery | 23 | 15.65 |
| No | Non-surgical procedure | 2 | 1.36 |
| No | Other reversal indication | 2 | 1.36 |
| Yes | Intracranial bleed – traumatic | 27 | 18.37 |
| Yes | Intracranial bleed – non-traumatic | 31 | 21.09 |

**Table 2.** Demographics and Patient Background

|  | Full Cohort<br>(N=147) | Non-ICH<br>(N=89) | ICH (N=58) | p-value |
| --- | --- | --- | --- | --- |
| Age, years (mean ± std.) | 73.0 ± 11.5 | 72.0 ± 11.7 | 74.5 ± 11.1 | 0.1812 |
| Male | 84 (57.1) | 53 (59.6) | 31 (53.4) | 0.4650 |
| Weight, kg (mean ± std.) | 91.3 ± 27.0 | 95.3 ± 29.6 | 85.1 ± 21.2 | 0.0157 |
| BMI, kg/m<sup>2</sup> (mean ± std.) | 31.3 ± 8.5 | 32.5 ± 9.1 | 29.4 ± 7.1 | 0.0344 |
| Caucasian (Non-Hispanic) | 137 (93.2) | 81 (91.0) | 56 (96.6) | 0.3161* |
| Medical History |  |  |  |  |
| CHF | 48 (32.7) | 27 (30.3) | 21 (36.2) | 0.4582 |
| Hypertension | 120 (81.6) | 70 (78.7) | 50 (86.2) | 0.2476 |
| CAD | 59 (40.1) | 32 (36.0) | 27 (46.6) | 0.2002 |
| MI | 18 (12.2) | 8 (9.0) | 10 (17.2) | 0.1357 |
| COPD | 35 (23.8) | 22 (24.7) | 13 (22.4) | 0.7484 |
| Hyperlipidemia | 95 (64.6) | 51 (57.3) | 44 (75.9) | 0.0214 |
| Diabetes Mellitus | 60 (40.8) | 35 (39.3) | 25 (43.1) | 0.6488 |
| Ischemic Stroke | 33 (22.4) | 19 (21.3) | 14 (24.1) | 0.6920 |
| Hemorrhagic Stroke | 13 (8.8) | 2 (2.2) | 11 (19.0) | 0.0005 |
| CKD | 39 (26.5) | 23 (25.8) | 16 (27.6) | 0.8150 |
| Cirrhosis | 1 (0.7) | 1 (1.1) | 0 (0.0) | 1.0000* |
| Vascular Disease | 14 (9.5) | 8 (9.0) | 6 (10.3) | 0.7843 |
| Factor Xa Inhibitor |  |  |  |  |
| Apixaban | 86 (58.5) | 54 (60.7) | 32 (55.2) | 0.5082 |
| Rivaroxaban | 61 (41.5) | 35 (39.3) | 26 (44.8) |  |
| Apixaban dose |  |  |  |  |
| 5 mg | 8 (9.4) | 7 (12.9) | 2 (6.2) | NA |
| 10 mg | 76 (89.4) | 46 (85.2) | 30 (93.8) |  |
| 20 mg | 1 (1.2) | 1 (1.9) | 0 (0.0) |  |
| Rivaroxaban dose |  |  |  |  |
| 10 mg | 1 (1.7) | 0 (0.0) | 1 (4.0) | NA |
| 15 mg | 10 (16.9) | 6 (17.7) | 4 (16.0) |  |
| 20 mg | 47 (79.7) | 27 (79.4) | 20 (80.0) |  |
| 30 mg | 1 (1.7) | 1 (2.9) | 0 (0.0) |  |
| Anticoagulant Indication |  |  |  |  |
| Atrial fibrillation | 131 (89.1) | 79 (88.8) | 52 (89.7) | 0.8654 |
| Venous Thromboembolism | 14 (9.5) | 9 (10.1) | 5 (8.6) | 0.7633 |
| VTE Prophylaxis | 6 (4.1) | 3 (3.4) | 3 (5.2) | 0.6807* |
| Other Indication | 4 (2.7) | 2 (2.3) | 2 (3.4) | 0.6472* |
| Admitting Service |  |  |  |  |
| Trauma | 35 (23.8) | 19 (21.3) | 16 (27.6) | <0.0001 |
| General Surgery | 21 (14.3) | 20 (22.5) | 1 (1.7) |  |
| Neurosurgery/Neurocritical Care | 38 (25.9) | 0 (0.0) | 38 (65.5) |  |
| Internal Medicine/Medical ICU | 43 (29.2) | 40 (44.9) | 3 (5.2) |  |
| Other Admitting Service | 10 (6.8) | 10 (11.2) | 0 (0.0) |  |
| Baseline Serum Creatinine, mg/dL<br>(mean ± std.) | 1.1 ± 0.5 | 1.2 ± 0.5 | 1.1 ± 0.5 | 0.2264 |
| Baseline Hemoglobin, g/dL (mean ±<br>std.) | 11.7 ± 2.8 | 10.9 ± 2.8 | 13.0 ± 2.4 | <0.0001 |
| Baseline GCS (1 not available) |  |  |  |  |
| 13-15 | 118 (80.8) | 77 (87.5) | 41 (70.7) | 0.0109 |
| 9-12 | 13 (8.9) | 3 (3.4) | 10 (17.2) |  |
| ≤8 | 15 (10.3) | 8 (9.1) | 7 (12.1) |  |
| FEIBA® dose, units (median [25<sup>th</sup>, 75<sup>th</sup><br>percentile]) | 4258 [3413, 4884] | 4254 [3440, 4900] | 4308 [3402, 4862] | 0.5776 |
| FEIBA Weight-Based Dose, units/kg<br>(median [25<sup>th</sup>, 75<sup>th</sup> percentile]) | 48.7 [45.5, 51.0] | 47.5 [44.4, 50.2] | 49.5 [47.2, 51.2] | 0.0155 |
| Additional Products |  |  |  |  |
| 2<sup>nd</sup> FEIBA Dose | 3 (2.0) | 3 (3.4) | 0 (0.0) | 0.2784* |
| Other Factor Products (Kcentra®,<br>NOVOSEVEN RT®, Profilnine®) | 5 (3.4) | 4 (4.5) | 1 (1.7) | 0.6485* |
| Anticoagulation Post-FEIBA® | 35 (23.8) | 34 (38.2) | 1 (1.7) | <0.0001 |
| Apixaban | 17 (48.6) | 16 (47.1) | 1 (100.0) | NA |
| Rivaroxaban | 13 (37.1) | 13 (38.2) | 0 (0.0) |  |
| Warfarin | 2 (5.7) | 2 (5.9) | 0 (0.0) |  |
| Other | 3 (8.6) | 3 (8.8) | 0 (0.0) |  |
| Time to Anticoagulation Resumption,<br>days (median [25<sup>th</sup>, 75<sup>th</sup> percentile]) | 78.5 [44.6, 116.0] | 77.7 [44.6, 111.1] | 125.2 | NA |
| VTE Prophylaxis Post-FEIBA® | 57 (38.8) | 23 (25.8) | 34 (58.6) | <0.0001 |
| Unfractionated Heparin | 45 (79.0) | 14 (60.9) | 9 (39.1) | NA |
| Enoxaparin | 12 (21.0) | 31 (91.2) | 3 (8.8) |  |
| Time to VTE Prophylaxis, days<br>(median [25<sup>th</sup>, 75<sup>th</sup> percentile]) | 34.1 [22.2, 52.4] | 22.2 [11.5, 45.1] | 41.7 [25.8, 72.5] | 0.0007 |
| Blood Products (PRBC and FFP) within<br>24 hours of FEIBA® (units) | 45 (30.6) | 41 (46.1) | 4 (6.9) | <0.0001 |
| Duration of Hospitalization, days<br>(median [25<sup>th</sup>, 75<sup>th</sup> percentile]) | 5.3 [3.0, 9.2] | 5.6 [3.2, 8.6] | 4.5 [2.6, 9.5] | 0.4361 |
| All numbers presented as N(%) unless otherwise noted |  |  |  |  |

**Table 3.** Description of Bleed

|  | Total (N=147) |
| --- | --- |
| ICH | N=58 |
| Subarachnoid | 15 (25.9) |
| Subdural | 22 (37.9) |
| Intraparenchymal | 21 (36.2) |
| Cerebellar | 7 (12.1) |
| Intraventricular | 6 (10.3) |
| Other | 1 (1.7) |
| Non-ICH | N=89 |
| Gastrointestinal | 27 (30.3) |
| Genitourinary | 1 (1.1) |
| Retroperitoneal | 3 (3.4) |
| Other Type of Bleed | 6 (6.7) |
| Emergency Surgery | 23 (25.8) |
| Trauma (Non-ICH) | 25 (28.1) |
| Non-Surgical Procedure | 2 (2.2) |
| Other Reversal Indication | 2 (2.2) |
| All numbers presented as N(%) |  |

**Table 4.** Effectiveness and Safety Results

|  | Full Cohort<br>(N=147) | Non-ICH<br>(N=89) | ICH<br>(N=58) | p-value |
| --- | --- | --- | --- | --- |
| Hemostasis |  |  |  |  |
| Excellent | 89 (60.5) | 46 (51.7) | 43 (74.1) | 0.0049* |
| Good | 26 (17.7) | 22 (24.7) | 4 (6.9) |  |
| Poor/None | 31 (21.1) | 21 (23.6) | 10 (17.2) |  |
| Unknown | 1 (0.7) | 0 (0.0) | 1 (1.7) |  |
| Hemostasis – Good or Excellent | 115 (78.2) | 68 (76.4) | 47 (81.0) | 0.5061 |
| Thrombotic Complication | 3 (2.0) | 2 (2.2) | 1 (1.7) | 1.0000* |
| Systemic Embolism | 1 | 1 | 0 |  |
| Ischemic Stroke | 1 | 0 | 1 |  |
| Other | 1 | 1 | 0 |  |
| Hemorrhagic Complication | 3 (2.0) | 3 (3.4) | 0 (0.0) | 0.2784* |
| Time Until Complication, Days (median [25<sup>th</sup>, 75<sup>th</sup> percentile]) | 1.4 [0.5, 2.9] | 1.1 [0.5, 2.9] | 1.7 | NA |
| In-hospital mortality | 15 (10.2) | 9 (10.1) | 6 (10.3) | 0.9637 |
| Time to death, days (median [25<sup>th</sup>, 75<sup>th</sup> percentile]) | 3.0 [0.9, 5.8] | 1.8 [0.9, 5.8] | 3.3 [1.7, 3.9] | 0.4437 |
| All numbers presented as N(%) unless otherwise noted |  |  |  |  |

**Table 5.**
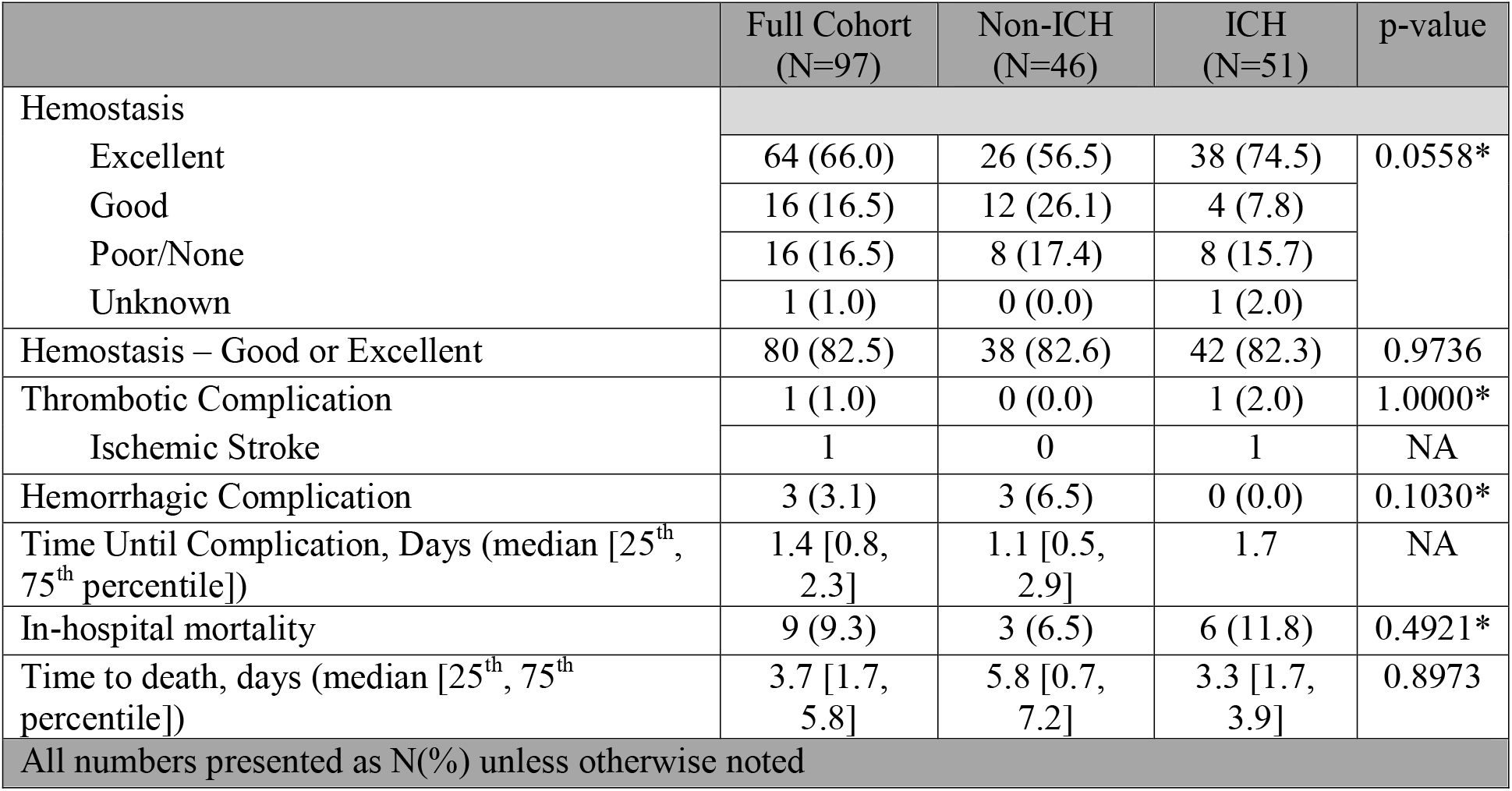
Effectiveness and Safety Results of Subgroup Paralleling ANNEXA-4 Trial^3^

## Appendix

**Appendix A.**
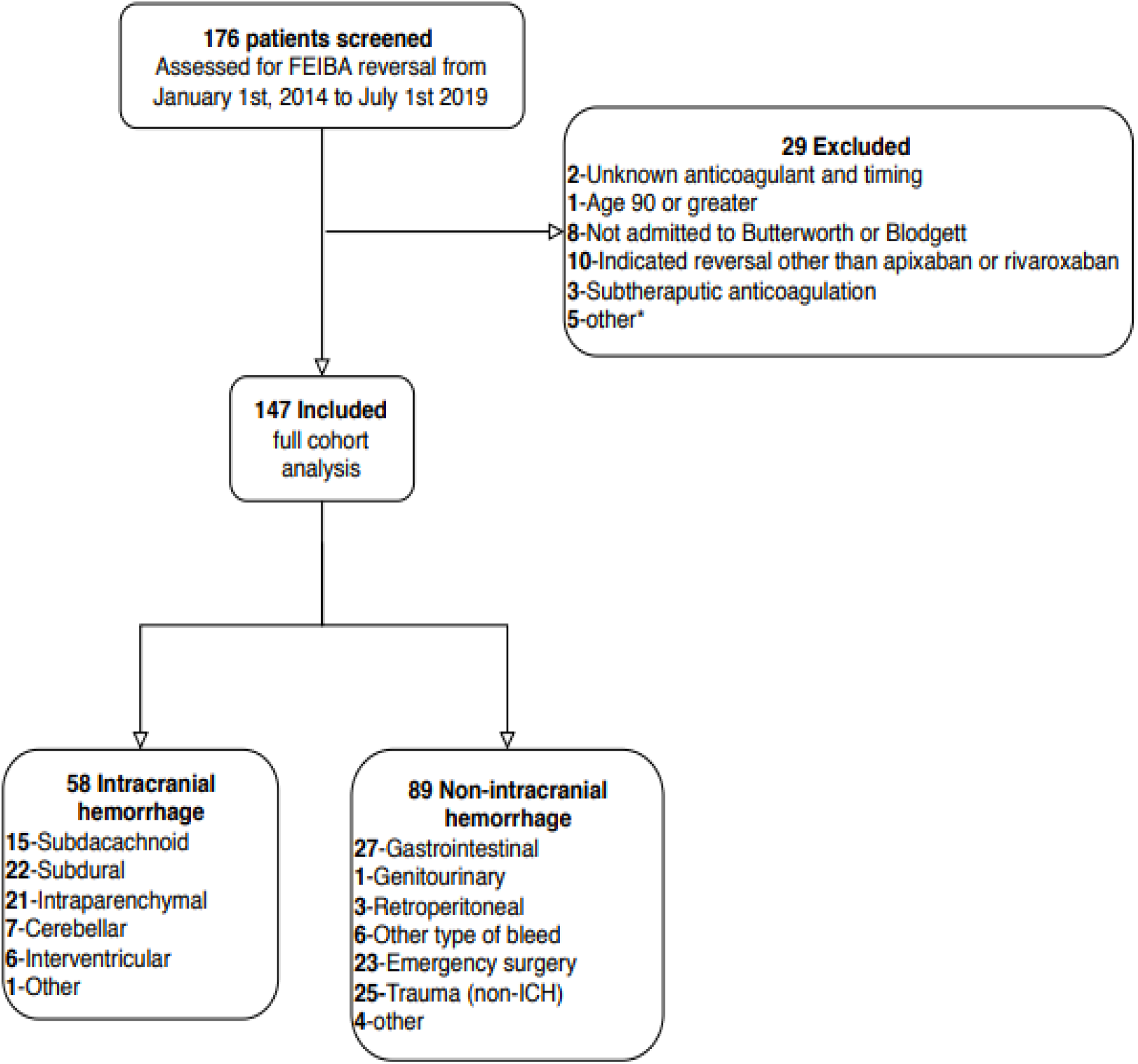
Inclusion Flowchart

**Appendix B.**
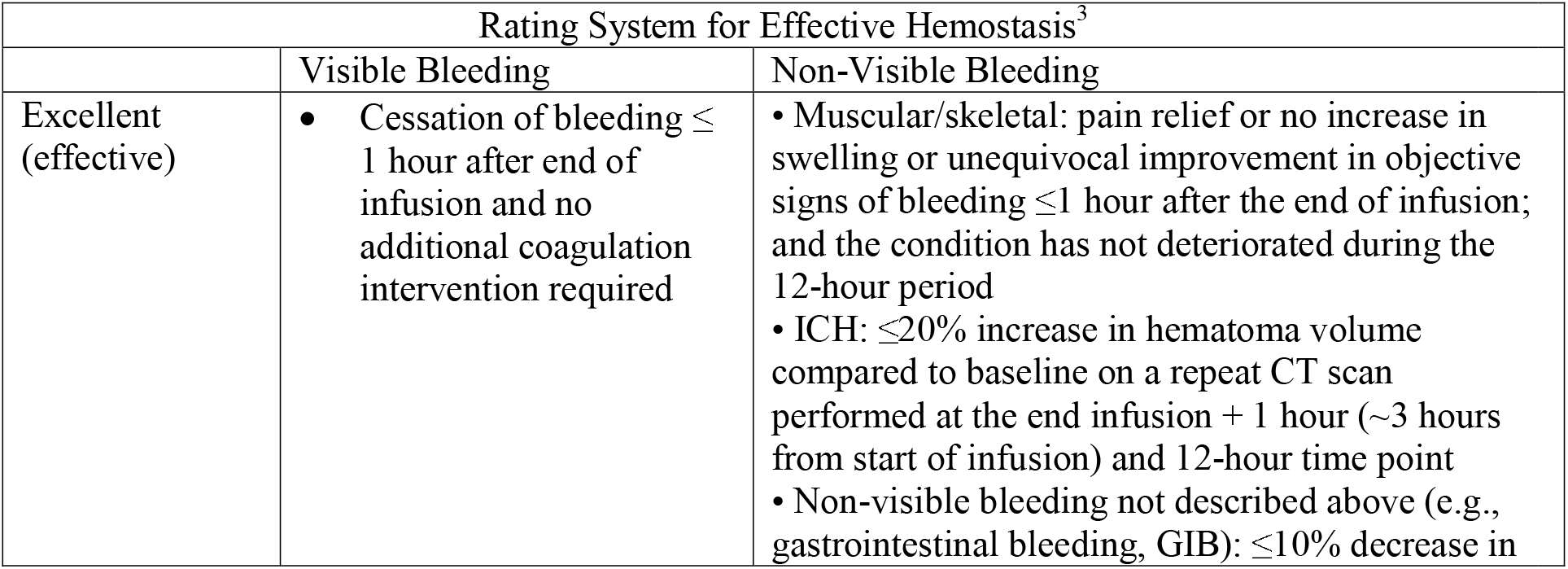
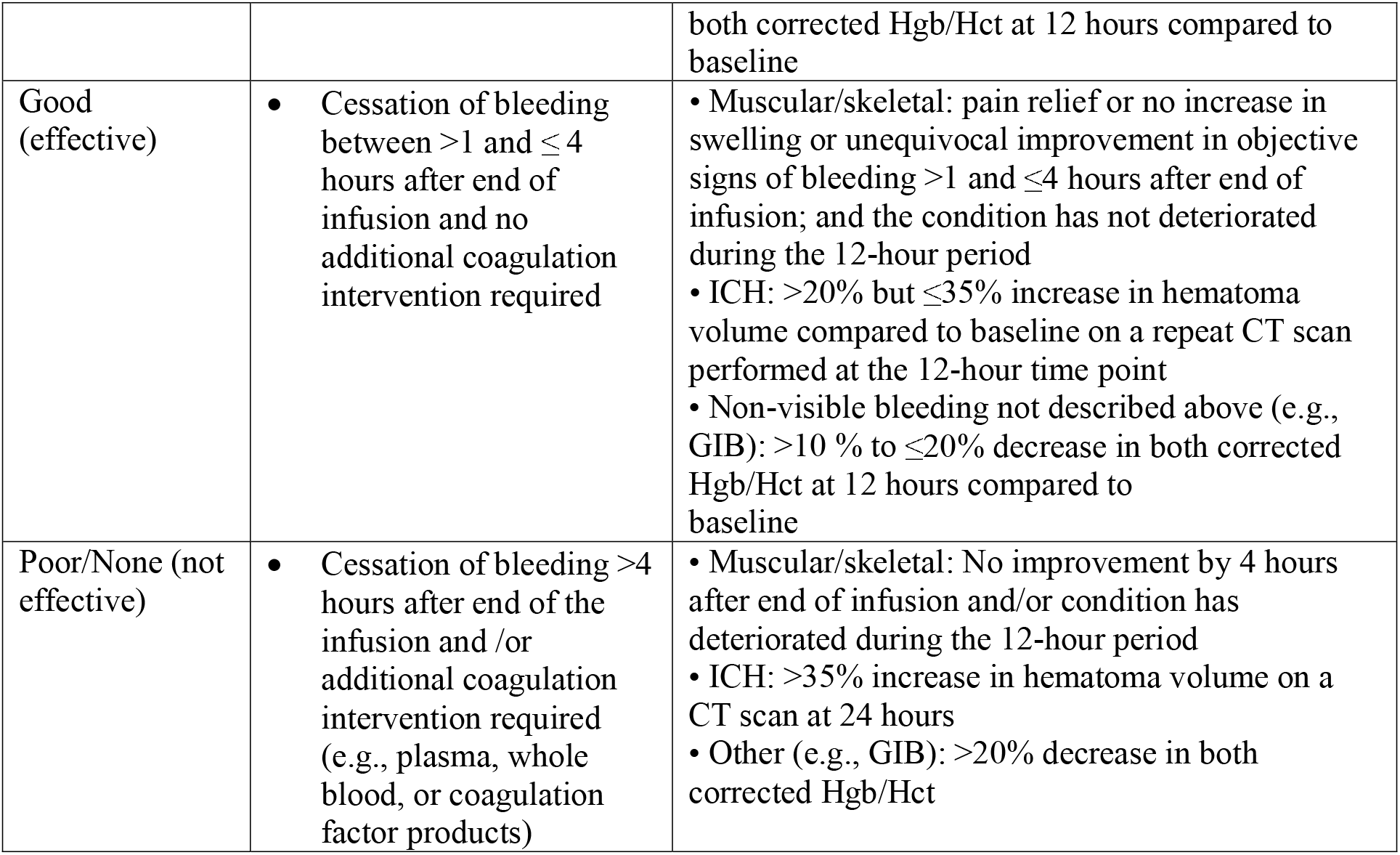
Hemostasis definition

## Notes

### Competing Interest Statement

The authors have declared no competing interest.

### Clinical Trial

Not registered since a retrospective study.

### Funding Statement

No funding was sought or obtained for this retrospective study.

### Author Declarations

Spectrum Health IRB & Ferris State University IRB.

